# Modern contraceptive Use and its associated factors among married women in Senegal: Based on the recent National Demographic and Health Survey Data

**DOI:** 10.1101/2022.01.30.22270102

**Authors:** Bezawit Mulat, Menen Tsegaw, Kegnie Shitu

## Abstract

**Objective:** This study aimed to assess the prevalence and associated factors of modern contraceptive use among married women in Senegal.

**Method:** The most recent Demographic and Health Survey (DHS), which was conducted in 2019 in Senegal was used. For this study 5,659 women were included. Multivariable binary Logistic regression analysis was used to identify factors associated with women’s contraceptive use.

**Result:** According to the study, 25.5% (95%CI (24.4, 26.7)) of married women in Senegal uses modern contraceptive methods. Being currently working (AOR=1.27, 95% CI (1.11, 1.46)), husband educational level (primary educational level (AOR=1.32, 95% CI (1.07,1.61), higher educational levels (AOR= 1.83, 95% CI (1.32, 2.54)), wealth index (middle (AOR=1.37,95% CI (1.15,1.63) and rich (AOR=1.34,95% CI (1.11,1.62)), exposure to family planning messages (AOR=1.29,95% CI (1.13,1.48)), having ANC visits(AOR=2.01, 95% CI(1.13,3.57)) and not wanting a child (AOR=2.35, 95%CI (1.90,2.95)) were positively and significantly with women’s contraceptives use. On the other hand, number of children (having no child (AOR=0.03, 95% CI (0.01,0.06)), 1-2 children (AOR=0.47,95% CI(0.38,0.59) and 3-4 children (AOR=0.79, 95% CI(0.67,0.96)), women who thought ≥5 children are ideal (AOR=0.73,95% CI(0.61,0.88)), and had a desire for more children after 2 years (AOR=2.19, 95% CI(1.82,2.64) were negatively and significantly with women’s contraceptives use.

**Conclusion:** contraceptive utilization was low among married women in Senegal. Women’s contraceptive use was affected by family planning messages exposure, sociodemographic, and obstetric factors. Thus, a tailored behavior change intervention is required to increase contraceptive utilization among married women in Senegal.

Strengths and limitations of this study

- The inclusion of nationally representative data and a large sample size, which strengthened the generalizability of the findings, was the study’s key strength.
- Sample weighing was employed to compensate unequal sample allocations and non-responses.
- It may not be possible to establish causal links due to the cross-sectional nature of the data.
- It’s possible that crucial factors that influence women’s use of modern contraceptives might be neglected because the study relied on secondary data.

## Introduction

Contraception today includes a variety of options: oral contraceptive pills, implants, injectable, contraceptive patch and vaginal ring, intrauterine device (IDU), female and male condoms, female and male sterilization, vaginal barrier methods (including the diaphragm, cervical cap and spermicidal agents), lactational amenorrhea method (LAM), emergency contraception pills, standard days method (SDM), basal body temperature (BBT) method, Two-day method and sympto-thermal method (1). Modern contraceptive use is characterized as a person’s or couple’s ability to have as many children as they want, when they want and has been found to lower mother and child morbidity and death (2,3).

The use of modern contraceptives is critical for achieving SDG 3’s goal 3.7, which states that by 2030, everyone in the world would have access to sexual and reproductive health care (4). Furthermore, in 1987, the Safe Motherhood Initiative established family planning as one of four pillars, alongside prenatal care, safe delivery, and postnatal care, to reduce maternal mortality in developing countries, where 99 percent of all maternal deaths occur (5).

According to the World Health Organization (WHO) report, nearly 308 million unwanted pregnancies were averted in 2017 (6). The increased use of contraceptive techniques has resulted in gains in not only health-related outcomes such as reduced maternal and newborn mortality, but also in educational and economic outcomes, particularly for girls and women (3,7).

In Africa the use of modern contraceptive varies across different countries. For instance, it is 62% in Zimbabwe, 6% in Guinea, 56%, Malawi and 56 % Kenya (8). Different studies stated that age, educational attainment of a woman, educational attainment of husbands, residence, wealth index of women, number of living children, having antenatal care visits (ANC), desire to space and want no more child are factors which have significant association with modern contraceptive utilization (8–10). Because of the large unmet need for contraception in Sub-Saharan African countries, the number of unplanned pregnancies is higher, estimated to be over 14 million per year (11). According to reports, 31% of conceptions in Senegal are unintentional, with 24 percent of all unwanted pregnancies ending in induced abortions, 60% in unplanned births, and 16% in miscarriages (12). All the aforementioned adverse outcomes would be prevented using contraceptives. However, there is a scarcity of data on the prevalence and associated aspects of modern contraception use in Senegal. As a result, the goal of this study is to determine the prevalence of modern contraceptive use and the factors that influence it among married women in Senegal.

## Methods

### Study setting, data source, and study design

Senegal is a West African country that borders the North Atlantic Ocean to the west. It is bordered on the north by Mauritania along the Senegal River, on the east by Mali, on the south by Guinea and Guinea-Bissau, and on the west by Mauritania along the Gambia River (13). Currently the population of the Senegal is estimated that 17,340,913 (14).

This analysis employed based on the data from the 2019 Continuous-Demographic and Health Survey (C-DHS) of Senegal. The C-DHS is a nationally representative data set developed by the Senegalese Agency of Statistics and Demography (ANSD) and the survey lasts from April 2019 - December 2019. The research participants are chosen using a two-stage stratified sampling technique. This data set (IR file) is consist of information collected from all eligible women aged 15-49 years and the current study excludes unmarried women and employed with a total weighted sample of 5,659 married reproductive age women. An authorization letter for the use of this data was obtained from DHS program and the dataset was downloaded from DHS website www.measuredhs.com

### Study variables

#### Dependent variable

The study’s outcome variable was the use of modern contraceptive methods among married reproductive age women in Senegal. The variable was catigorized into two categories: 1 = “Use modern contraceptive” and 0 =“did not use modern contraceptive”.

#### Independent variables

In this study the independent variables included: sociodemographic factors (Age, religion, residence, educational status of women, educational status of husband, and current working (employment) status, wealth index, media exposure to family planning methods and obstetric factors: complications during previous pregnancies (terminated pregnancy), presence of antenatal care visits, number of living children, ideal number of children that the family want and desire for more children.

#### Data processing and Analysis

Individual records (IR) files were used to extract data, which was then coded and transformed using STATA version 14 statistical software. To account for differential chance of selection and non-response in the original survey, weighted samples were used for analysis. The presence of statistical significance was determined using a multivariable logistic regression analysis. It was fitted after the model’s fitness was evaluated using the Hosmer and Lemeshow goodness of fit test. The variance inflation factor (VIF) was also used to analyze multicollinearity across the explanatory components, and it was found to be within acceptable range (1-5)(15). A p-value less than 0.05 is used to evaluate the presence of a meaningful effect or relationship of independent factors with the outcome variable.

#### Patient public involvement

The public was not involved in the design or conduct, choice of outcome measures and recruitment of the study.

## Result

A total of 5,659 married women were included in the study. The mean age of the participants was 31 (± 8.5) years with an age range of 15-49 years. The majority (98.2%) of the mothers were Muslims and 3357 (59.3 %) were from a rural area. Concerning the reproductive history (98.6%) of women have ANC visits (table 1).

**Table 1.**
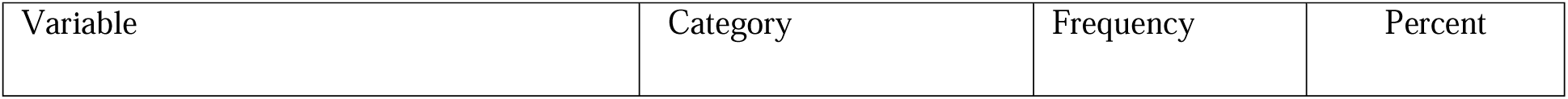
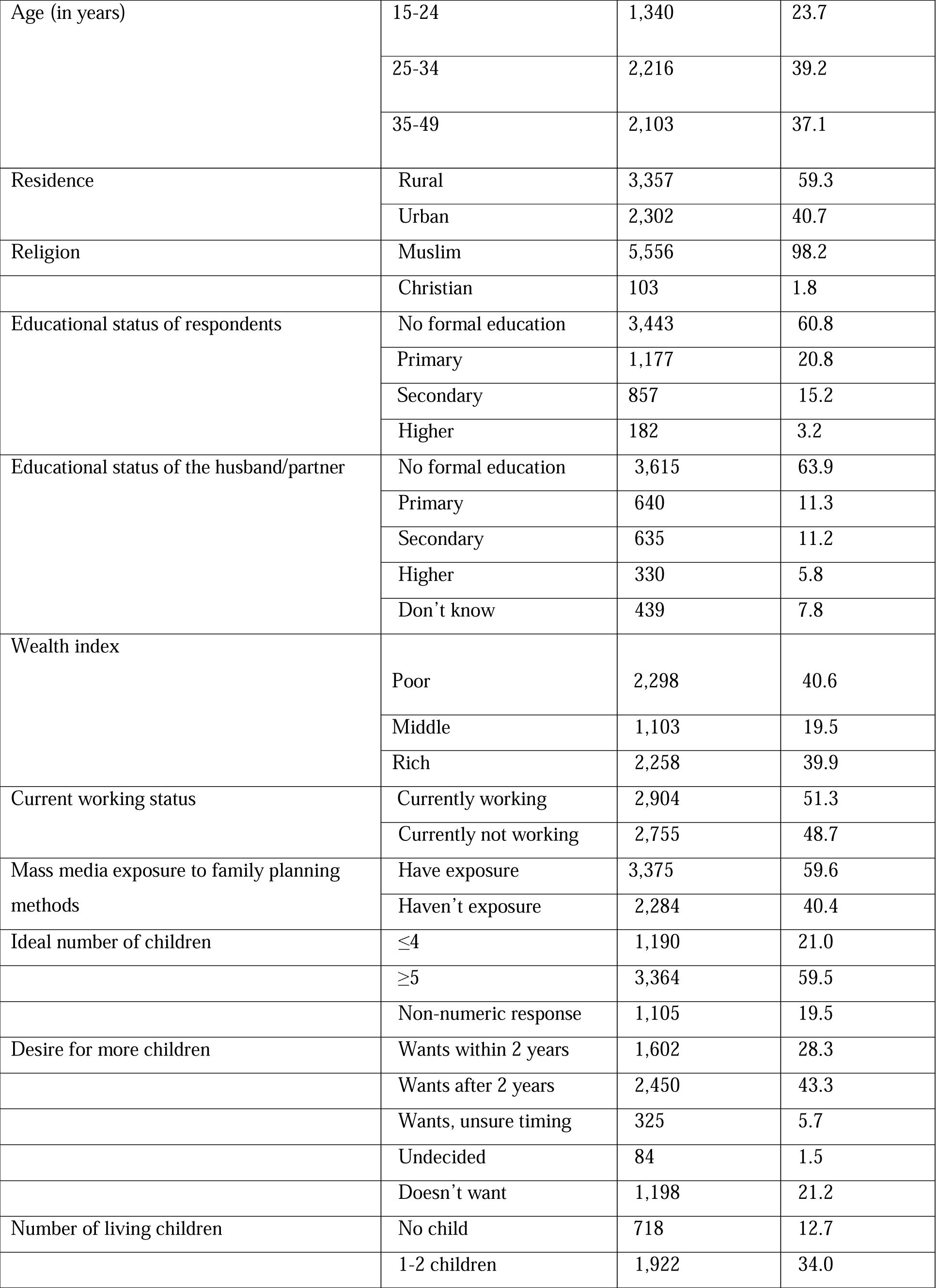
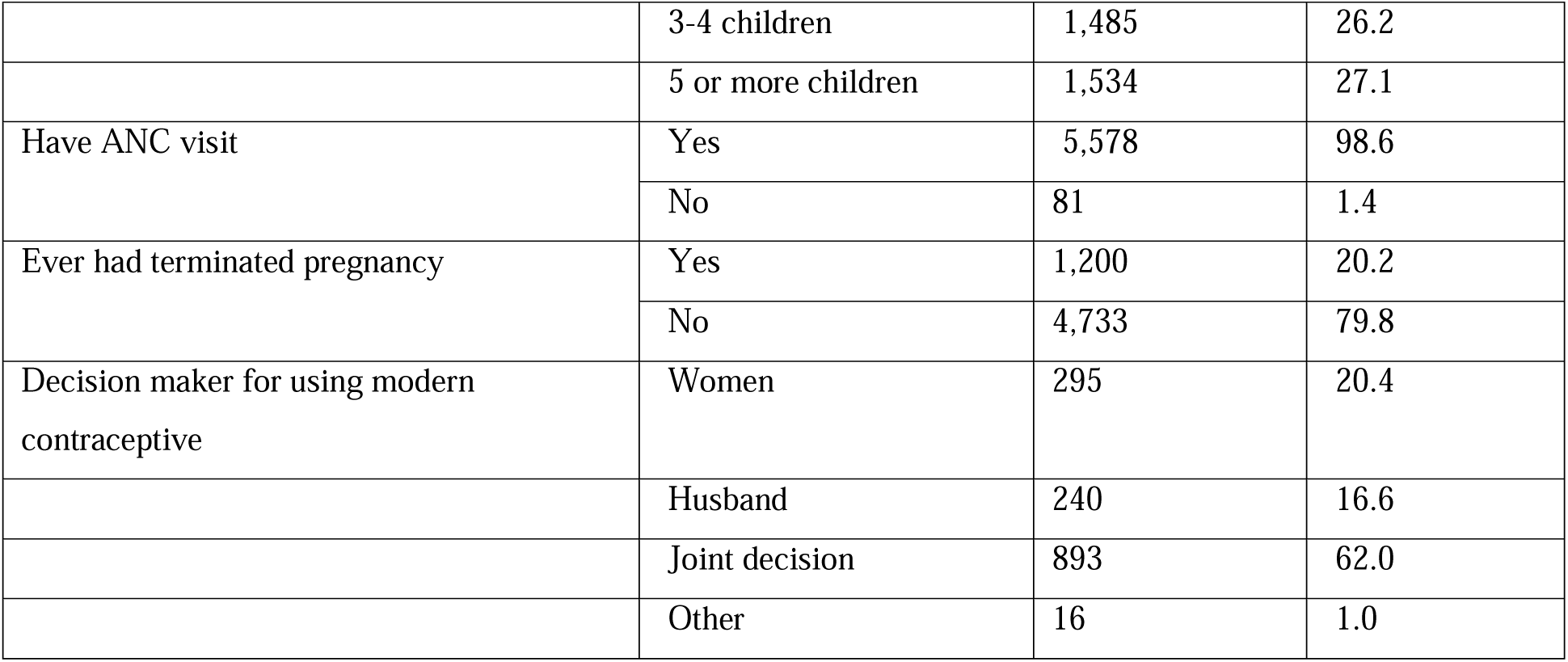
Sociodemographic and reproductive characteristics of married women in Senegal (n = 5,659).

### Modern contraceptive use among married women

In this study the prevalence of modern contraceptive use among married women in Senegal was 25.5% with (95% CI (24.4, 26.7)). The use of modern contraceptive method is higher among age group of 25-34 as compared to age group of women 15-24. Additionally, the utilization of modern contraceptives was higher (30.5%) among women who are currently working. Furthermore, women who are exposed to family planning methods through the media are more likely to use modern contraception (29.5%).

### Factors associated with modern contraceptive use

Based on the output of multivariable binary logistic regression analysis, the following explanatory variables had a statistically significant association with modern contraceptive use : current working status of women(currently working (AOR=1.27, 95% CI (1.11, 1.46))), Husband education (husbands who attended primary educational level (AOR=1.32, 95% CI (1.07,1.61), husbands who attended higher educational levels (AOR= 1.83, 95% CI (1.32, 2.54), women whose wealth index in middle(AOR=1.37,95% CI (1.15,1.63) and rich (AOR=1.34,95% CI (1.11,1.62), women who had exposed to family planning methods through different medias (AOR=1.29,95% CI (1.13,1.48)), women who had ANC visits(AOR=2.01, 95% CI(1.13,3.57)). Plus to the above factors, women who had no child (AOR=0.03, 95% CI (0.01,0.06)), 1-2 children (AOR=0.47,95% CI(0.38,0.59) and 3-4 children (AOR=0.79, 95% CI(0.67,0.96)), women who thought ≥5 children are ideal(AOR=0.73,95% CI(0.61,0.88)), and women who had a desire for more children after 2 years (AOR=2.19, 95% CI(1.82,2.64) and women who doesn’t want a child (AOR=2.35, 95%CI (1.90,2.95)) were significantly associated with womens modern contraceptive use.

The odds of using modern contraceptive was increased by 27% among women who were currently working than their counterparts. The odds of using modern contraceptives was increased by 32% among women whose husband had primary level of education. Moreover, having a husband with primary and higher education level increased the odds of using modern contraceptive by 32% and 16% respectively, when compared to women having an illiterate husband. The odds of using modern contraceptives was increased by 29% among women who had exposure to family planning methods through different Medias (Table 2).

**Table 2.** Factors associated with modern contraceptive use among married women in Senegal (n=5,659).

| Variable | Modern contraceptive use |  | p-value | AOR |
| --- | --- | --- | --- | --- |
|  | Yes (n=1,445) | No(n=4,214) |  |  |
|  | Frequency (%) | Frequency (%) |  |  |
| **Age** |  |  |  |  |
| 15-24 | 221(16.5) | 1,119(83.5) |  | 1 |
| 25-34 | 613(27.7) | 1,603(72.3) | 0.8 | 0.97(0.78,1.2) |
| 35-49 | 611(29.1) | 1,492(70.9) | 0.14 | 0.82(0.64,1.1) |
| **Residence** |  |  |  |  |
| Urban | 745(32.4) | 1,557(67.6) |  | 1 |
| Rural | 700(20.9) | 2,657(79.1) | 0.06 | 0.85(0.72,1.00) |
| **Religion** |  |  |  |  |
| Muslim | 1,414(25.5) | 4,142(74.5) |  | 1 |
| Christian | 31 (29.7) | 72 (70.3) | 0.92 | 0.98 (0.62,1.55) |
| **Womens' educational level** |  |  |  |  |
| No formal education | 754(21.9) | 2,690(78.1) | 0.05 | 0.61(0.37,1.00) |
| Primary | 374(31.8) | 803(68.2) | 0.58 | 0.87(0.53,1.42) |
| Secondary | 240(28.0) | 617(72.0) | 0.54 | 0.86(0.53,1.40) |
| Higher | 77(42.3) | 105(57.7) |  | 1 |
| **Current working status** |  |  |  |  |
| Currently working | 841(30.5) | 1,913(69.5) | 0.00 | 1.27 (1.11,1.46) |
| Currently not working | 604(20.8) | 2,301(79.2) |  | 1 |
| **Husband education** |  |  |  |  |
| No formal education | 777(21.5) | 2838(78.5) |  | 1 |
| Primary education | 192(29.9) | 448(70.1) | 0.008 | 1.32 (1.07,1.61) |
| Secondary education | 192(30.2) | 443(69.8%) | 0.19 | 1.16 (0.93,1.45) |
| Higher | 151(45.9) | 179(54.1) | 0.00 | 1.83 (1.32,2.54) |
| Don't know | 134(30.4) | 306(69.6) | 0.28 | 1.16 (0.89,1.51) |
| **Wealth index** |  |  |  |  |
| Poor | 434(18.9) | 1863(81.1) |  | 1 |
| Medium | 301(27.3) | 802(72.7) | 0.00 | 1.37 (1.15,1.63) |
| Rich | 709(31.4) | 1549(68.6) | 0.00 | 1.34 (1.11,1.62) |
| **Media exposure** |  |  |  |  |
| Exposed | 995(29.5) | 2,381(70.5) | 0.00 | 1.29 (1.13,1.48) |
| Non exposed | 450(19.7) | 1,833(80.3) |  | 1 |
| **Have ANC visits** |  |  |  |  |
| Yes | 1436(25.8) | 4,142(74.5) | 0.01 | 2.01 (1.13,3.57) |
| No | 9(10.7) | 72(89.3) |  | 1 |
| **Ever had terminated pregnancy** |  |  |  |  |
| Yes | 312(26.3) | 877(73.7) | 0.41 | 0.94 (0.79,1.09) |
| No | 1,132(25.3) | 3,337(74.7) |  | 1 |
| **Number of living children** |  |  |  |  |
| No children | 8(1.1) | 711(98.9) | 0.00 | 0.03 (0.01,0.06) |
| 1-2 children | 474(24.7) | 1,448(75.3) | 0.00 | 0.47 (0.38,0.59) |
| 3-4 children | 470(31.7) | 1,014(68.3) | 0.02 | 0.79 (0.67,0.96) |
| ≥5 children | 493(32.1) | 1,041(67.9) |  | 1 |
| **Ideal number of children** |  |  |  |  |
| ≤ 4 children | 381(32.0) | 809(68.0) |  | 1 |
| ≥ 5 children | 854(25.4) | 2,509(74.6) | 0.001 | 0.73(0.61,0.88) |
| Non-numeric response | 210(19.0) | 895(81.0) | 0.00 | 0.52(0.42,0.66) |
| **Desire for more children** |  |  |  |  |
| Want's within 2 years | 195(12.2) | 1,407(87.8) |  | 1 |
| Want's after 2 years | 750(30.6) | 1,700(69.4) | 0.00 | 2.19(1.82,2.64) |
| Want's, unsure timing | 37(11.4) | 288(88.6) | 0.49 | 1.13(0.79,1.60) |
| Undecided | 14(17.4) | 70(82.6) | 0.40 | 1.27(0.72,2.24) |
| Doesn't want | 447(37.4) | 750(62.6) | 0.00 | 2.35(1.90,2.95) |

## Discussion

This study assessed the prevalence and associated factors of modern contraceptive use among Senegalese women. The prevalence of modern contraceptive use was 25.5% with (95% CI (24.4, 26.7)). Womens’ working status, husband education, wealth index, having media exposure, having ANC visit, number of living children, ideal number of children and desire for more children were factors that have statistically significant association with modern contraceptive use.

In the present study, the use of modern contraceptives among married Senegalese women was 25.5% (95% CI (24.4, 26.7). The result is in harmony with the study conducted in Senegal (26.3%) (16). This alignment might be explained by similarities in sociodemographic characteristics of the population.

The result of the current study is higher than the studies conducted in Bale zone, south East Ethiopia (20.8%)(17) and Metekel zone North West Ethiopia (18.6%)(18). The discrepancy might be explained by socio-cultural difference between the countries that could affect their awareness on contraceptive use (19). On the other hand the result of the present study is lower than studies done in North Showa, Ethiopia (46.9%) (20) Zambia (43%)(21), and Egypt (46.5%) (22).

In the present study women who are currently enrolled in different jobs (currently working) had significant association with modern contraceptive use. This result is supported by study conducted in Ethiopia (19). This could be owing to the fact that women who work and have a private job have a higher income, greater access to the media, and better health care, all of which have a beneficial impact on modern contraception use (23–26).

In the current study, modern contraceptive use among married Senegalese women was significantly associated with husband’s educational attainment, which is supported by a study conducted in Senegal (16,27,28). It’s probable that educated husbands are aware of the benefits of modern contraception since they have a greater chance to read newspapers, watch television, and use social media. The other factor that had significant association with contraceptive use was wealth index of a woman. Women from households with a medium or high wealth index were more likely to use modern contraception than those from households with a low wealth index. The finding was in line with a study conducted in Ethiopia (29).

Modern contraceptive use was also significantly associated with womens’ media exposure. This outcome is obedient with studies conducted in Senegal (16) and Ethiopia (9). This alignment could be explained by the fact that media can help communities raise awareness about health issues as well as address social and cultural issues (30,31). This critical role of the media can aid in lowering barriers to health-care access and use, including contraception.

In this study presence of ANC visits have significant association with modern contraceptive use. The result is supported by a multicenter study done in African countries (8). It’s possible that the increased prevalence of contraceptive use among women who had ANC visits is due to the fact that antenatal care provides a unique opportunity to interact with, educate, and provide family support to women of all socioeconomic backgrounds, which facilitates the uptake of modern contraceptives. The study also showed that women who hadn’t child have lower tendency to use modern contraceptives than women who had three and more than three children. This result is supported by studies done in Senegal (16,32). This meant that the more children a woman has, the more likely she is to want to space or limit the number of children, and the more contraceptive she is likely to use.

Furthermore, the present study also revealed the presence of statistically significant association between modern contraceptive use, and womens’ perception for ideal number of children they would have. The result is in harmony with a study done in Senegal (16). According to the study, women who wished five or more children had a lower likelihood of taking modern contraception than women who desired four or fewer children. Finally, the results of the current study also showed that womens’ desire for more children also affects womens’ modern contraceptive utilization. That is the odds of using modern contraceptives among women who wants child within 2 years lower than women who wants child after 2 years and totally doesn’t want a child at all.

The study’s main strength was the use of nationally representative data and a large sample size, which increased the generalizability of its conclusions. The DHS study, on the other hand, has flaws in that it relies on respondents’ self-reporting and may be prone to recall bias because women were asked to recall past incidents. Furthermore, the design of this study limits its ability to demonstrate causation between the desired outcome and these critical independent factors.

### Conclusion and recommendation

According to the findings, the usage of modern contraceptive methods among married women in Senegal was low. Women’s contraceptive use was affected by family planning messages exposure, sociodemographic, and obstetric factors. Thus, a tailored behavior change intervention is required to increase contraceptive utilization among married women in Senegal and such interventions should take those factors into account.

## Data Availability

The data used to analyse for the current study is publicly available with data archivist request approval.

https://www.measuredhs.com

## Ethics approval and consent to participate

The study is based on DHS data which was collected by DHS program, the DHS program collect the data by gaining ethical approval from national IRB. Thus, we didn’t received any ethical approval since the data used for the study is from a secondary source. However, we got the authentication letter from the DHS program to use the data set for this study. The study was conducted under the Declaration of Helsinki.

## Consent for publication

Not applicable.

## Availability of data and materials

All result-based data are available within the manuscript and anyone can access the data set online from www.measuredhs.com

## Authors’ contributions

All authors made substantial contributions to conception, acquisition of data, or analysis and interpretation of data; took part in drafting the article or revising it critically for important intellectual content; agreed to submit to the current journal; gave final approval of the version to be published; and agree to be accountable for all aspects of the work.

## Acknowledgments

We would like to acknowledge the MEASURE DHS program for permitting us to obtain and use DHS data sets of Senegal.

